# Risk of acute arterial and venous thromboembolic events in Eosinophilic Granulomatosis with Polyangiitis (Churg-Strauss syndrome)

**DOI:** 10.1101/2020.11.09.20228197

**Authors:** Alessandra Bettiol, Renato Alberto Sinico, Franco Schiavon, Sara Monti, Enrica Paola Bozzolo, Franco Franceschini, Marcello Govoni, Claudio Lunardi, Giuseppe Guida, Giuseppe Lopalco, Giuseppe Paolazzi, Angelo Vacca, Gina Gregorini, Pietro Leccese, Matteo Piga, Fabrizio Conti, Paolo Fraticelli, Luca Quartuccio, Federico Alberici, Carlo Salvarani, Silvano Bettio, Simone Negrini, Carlo Selmi, Savino Sciascia, Gabriella Moroni, Loredana Colla, Carlo Manno, Maria Letizia Urban, Alfredo Vannacci, Maria Rosa Pozzi, Paolo Fabbrini, Stefano Polti, Mara Felicetti, Maria Rita Marchi, Roberto Padoan, Paolo Delvino, Roberto Caporali, Carlomaurizio Montecucco, Lorenzo Dagna, Adriana Cariddi, Paola Toniati, Silvia Tamanini, Federica Furini, Alessandra Bortoluzzi, Elisa Tinazzi, Lorenzo Delfino, Iuliana Badiu, Giovanni Rolla, Vincenzo Venerito, Florenzo Iannone, Alvise Berti, Roberto Bortolotti, Vito Racanelli, Guido Jeannin, Angela Padula, Alberto Cauli, Roberta Priori, Armando Gabrielli, Milena Bond, Martina Tedesco, Giulia Pazzola, Paola Tomietto, Marco Pellecchio, Chiara Marvisi, Federica Maritati, Alessandra Palmisano, Christian Dejaco, Johann Willeit, Stefan Kiechl, Iacopo Olivotto, Peter Willeit, Domenico Prisco, Augusto Vaglio, Giacomo Emmi, on behalf of the Italian EGPA Consortium

**Affiliations:** Department of Experimental and Clinical Medicine, University of Firenze, Italy; Department of Neurosciences, Psychology, Drug Research and Child Health (NEUROFARBA), University of Firenze, Italy; Department of Medicine and Surgery, University of Milano – Bicocca and Nephrology Unit, ASST-Monza, Milan/Monza, Italy; Operative Unit of Rheumatology, Department of Medicine DIMED, University Hospital of Padova, Italy; Division of Rheumatology IRCCS Policlinico S.Matteo Foundation and University of Pavia, Pavia, Italy; University of Pavia, PhD in Experimental Medicine, Pavia, Italy; Unit of Immunology, Rheumatology, Allergy and Rare Diseases (UnIRAR), IRCCS-San Raffaele Scientific Institute, Milan, Italy; Unit of Rheumatology and Clinical Immunology, University and ASST Spedali Civili, Brescia, Italy; Department of Medical Sciences, University of Ferrara, Italy; Rheumatology Unit - Azienda Ospedaliero-Universitaria S. Anna - Ferrara (Cona); Department of Medicine, University of Verona, Verona, Italy; Allergy and Pneumology Unit, A.O. S. Croce e Carle, Cuneo, Italy; Rheumatology Unit, Department of Emergency and Organ Transplantation (DETO), Polyclinic Hospital, University of Bari, Italy; Department of Rheumatology, Santa Chiara Hospital, Trento, Italy; Department of Biomedical Sciences and Human Oncology, Unit of Internal Medicine “Guido Baccelli”, University of Bari “Aldo Moro” Medical School, Bari, Italy; Nephrology Unit, ASST Spedali Civili, Brescia, Italy; Rheumatology Department of Lucania - San Carlo Hospital, Potenza, Italy; Rheumatology, Department of Medical Sciences and Public Health, University Clinic, Cagliari, Italy; Rheumatology Unit, Department of Clinical Internal, Anesthesiological and Cardiovascular Sciences, Sapienza University of Rome, Rome, Italy; Department of Internal Medicine, Clinica Medica, Ospedali Riuniti, Ancona; Rheumatology Clinic, Department of Medicine (DAME), University of Udine, Udine, Italy; Department of Medical and Surgical Specialties, Radiological Sciences and Public Health, University of Brescia, Brescia, Italy; Nephrology Unit, Spedali Civili Hospital, Azienda Socio Sanitaria Territoriale degli Spedali Civili di Brescia, Brescia, Italy; Azienda USL-IRCCS di Reggio Emilia and University of Modena and Reggio Emilia; Rheumatology Unit, Internal Medicine Department, Cattinara Teaching Hospital (ASUITS) Trieste; Internal Medicine, Clinical Immunology and Translational Medicine Unit, IRCCS Ospedale Policlinico San Martino, Genoa, Italy; Centre of Excellence for Biomedical Research and Department of Internal Medicine, University of Genoa, Genoa, Italy; Division of Rheumatology and Clinical Immunology, Humanitas Clinical and Research Center IRCCS, Rozzano - Milan, Italy; Department of Biomedical Sciences, Humanitas University, Pieve Emanuele - Milan, Italy; Center of Research of Immunopathology and Rare Diseases- Coordinating Center of the Network for Rare Diseases of Piedmont and Aosta Valley, Department of Clinical and Biological Sciences, University of Turin, Italy; SCU Nephrology and Dialysis, S. Giovanni Bosco Hospital, Department of Clinical and Biological Sciences, University of Turin, Italy; Nephrology Unit, Fondazione IRCCS Ca’ Granda Ospedale Maggiore Policlinico, Milan, Italy; Nephrology, Dialysis and Renal Transplant Division, Department of Medical Sciences, “Città della Salute e della Scienza di Torino” University Hospital, University of Turin, Italy; Department of Emergency and Organ Transplantation, Nephrology, Dialysis and Transplant Unit, University of Bari “Aldo Moro”, Bari, Italy; Nephrology Unit, Hospital San Gerardo Monza, University of Milano Bicocca; Respiratory Pathophysiology Division, University Hospital of Padova, Italy; Division of Clinical Rheumatology, ASST Gaetano Pini-CTO Institute, Milan, Italy; Department of Clinical Sciences & Community Health, Research Center for Adult and Pediatric Rheumatic Diseases, Università degli Studi di Milano, Milan, Italy; Vita-Salute San Raffaele University, Milan, Italy; Department of Medical Science, Allergy and Clinical Immunology, University of Torino and AO Ordine Mauriziano Umberto I, Turin, Italy; Department of Internal Medicine, Università Politecnica delle Marche, Ancona, Italy; Department of Biomedical and Clinical Sciences “L. Sacco,” University of Milan, Milan - Italy; Struttura Complessa Medicina Interna 1 P.O. Levante - ASL 2 Savona Liguria; Nephrology Unit, University Hospital, Parma, Italy; Department of Experimental Diagnostic and Specialty Medicine (DIMES), Nephrology, Dialysis and Renal Transplant Unit, St. Orsola Hospital, University of Bologna, Bologna, Italy; Hospital of Brunico (SABES-ASDAA), Department of Rheumatology, Brunico, Italy; Department of Rheumatology and Immunology, Medical University Graz, Graz, Austria; Department of Neurology, Medical University of Innsbruck, Innsbruck, Austria; Cardiomyopathy Unit, Careggi University Hospital, Florence, Italy; Department of Public Health and Primary Care, University of Cambridge, Cambridge, UK; Nephrology and Dialysis Unit, Meyer Children’s Hospital, Florence, Italy; Department of Experimental, Clinical and Biomedical Sciences “Mario Serio”, University of Firenze, Florence, Italy

**Author notes:** **Address for correspondence:** Alessandra Bettiol, MSc PhD, Department of Experimental and Clinical Medicine, University of Firenze Largo Brambilla 3, 50134 Firenze, ITALY. These authors contributed equally to this manuscript (Peter Willeit, Domenico Prisco, Augusto Vaglio, Giacomo Emmi, on behalf of the Italian EGPA Consortium). **Conflict of interest statement:** Dr. Silvia Tamanini worked at ASST Spedali Civili Brescia, Unit of Rheumatology and Immunology at the moment of the study. At the moment of publication, she works at Glaxo Smith Kline. All other authors report no conflicts of interest. **Funding:**This study was not funded.

**Keywords:** Eosinophilic Granulomatosis with Polyangiitis, ANCA-associated Vasculitis, Thrombosis, Cardiovascular, Eosinophils

## Abstract

**Background and objective:** Systemic small vessel vasculitides carry an increased risk of acute arterial and venous thromboembolic events (AVTE); however, this risk has not been systematically explored in Eosinophilic Granulomatosis with Polyangiitis (EGPA). This study assessed the occurrence and main risk factors of AVTE among EGPA patients as compared to the general community from the population-based Bruneck cohort.

**Methods:** We conducted a retrospective multicenter cohort study on 573 EGPA patients. Clinical and serological data were collected at diagnosis. Occurrence of AVTE and time to the first AVTE after EGPA diagnosis were recorded. Age-standardized event rate (SER) of AVTE as compared to the reference cohort was assessed. Cox regression was applied to identify AVTE predictors.

**Results:** 129 EGPA patients (22.5%) had AVTE, considered as potentially life-threatening in 55.8%. Seventy patients experienced an AVTE prior to diagnosis (of whom 58.6% in the two years before diagnosis) and 75 following EGPA diagnosis, of whom 56% in the two subsequent years. The SER of AVTE as compared to the reference cohort was 2.10 (95% CI 1.67-2.63). This risk was particularly increased in patients with history of AVTE and with a Birmingham Vasculitis Activity Score ≥20 at diagnosis. Patients receiving immunosuppression within 2 months of diagnosis were at lower risk, while antiplatelet or anticoagulant treatment did not confer measurable benefit.

**Conclusion:** EGPA is associated with AVTE in approximately one quarter of patients, particularly around diagnosis. Immunosuppressants seemed to exert a protective effect, while anticoagulant and antiplatelet agents did not.

## INTRODUCTION

Eosinophilic Granulomatosis with Polyangiitis (EGPA, Churg-Strauss syndrome) belongs to the anti-neutrophil cytoplasmic antibody (ANCA)-associated vasculitides (AAVs), which also include Granulomatosis with Polyangiitis and Microscopic Polyangiitis[1, 2]. An increased frequency of vascular events due to the involvement of medium-sized and large vessels has been reported consistently in AAV[3–5]. Recent evidence indicates an association between disease activity and thrombotic risk in AAV regardless of clinical diagnosis or ANCA specificity, suggesting that vascular inflammation *per se* is a major determinant of thrombosis [6, 7].

EGPA is characterized by eosinophilic vasculitis and eosinophil-rich extravascular inflammation and typically presents with peripheral eosinophilia[8, 9]. Cardiac manifestations are reported in 40-60% of patients, and represent the leading cause of mortality[10, 11]. Recent reports suggest that acute arterial and venous thromboembolic events (AVTE) might represent a consistent burden of disease in EGPA patients[5], possibly due to vascular inflammation or to cardio-embolic events secondary to EGPA-related cardiomyopathy. Additionally, eosinophilia is a major determinant of thrombosis, as suggested by the notion that different eosinophilic conditions show a high prevalence of AVTE [12]. In the present study, we assessed incidence, type and timing of AVTEs in the largest EGPA cohort reported thus far, and compared incidence estimates with those in the general community obtained from the population-based Bruneck study cohort, and identified independent predictors of risk. We considered AVTEs that occurred both before and after EGPA diagnosis, and analyzed possible associations between AVTE, disease activity, other disease-related parameters and pharmacological treatments including anticoagulant and antiplatelet agents.

## MATERIALS AND METHODS

### Study design and setting

We conducted a retrospective observational cohort study. The study involved 28 Italian EGPA referral centres, including rheumatology, nephrology, clinical immunology, pulmonology and internal medicine units. It was conducted in compliance with the Declaration of Helsinki and approved by the Ethic Committee of the University Hospital Careggi, Florence, Italy (Approval number 12804/CAM_BIO).

### Participants

We consecutively included all EGPA patients followed between January 1988 and September 2018. The patients’ demographic, serological and clinical data were retrieved from medical charts and recorded into an electronic database created *ad hoc* for the study. All patients had to fulfil the American College of Rheumatology classification criteria for EGPA [13] or the criteria proposed in the MIRRA trial [14].

### Exposure and outcome variables

The following demographic and clinical data were collected at the time of EGPA diagnosis: age, gender, previous AVTEs and presence of cardiovascular risk factors and/or comorbidities (including smoking habit, hypercholesterolemia, hypertension, diabetes mellitus, and history of cancer) and ongoing cardiovascular therapies.

EGPA-related clinical and serological data at diagnosis were recorded, including specific organ manifestations, disease activity based on the Birmingham Vasculitis Activity Score (BVAS)[15], presence of ANCA, and laboratory tests including C-reactive protein, erythrocyte sedimentation rate, eosinophil count, eosinophil cationic protein, IgG4 and IgE titers. For each patient, the revised five-factor score was calculated [16], with one point each attributed to the following factors: cardiac, gastrointestinal or renal involvement; age >65 years; absence of ear-nose-throat involvement.

The immunosuppressive treatments started after diagnosis were also recorded. After EGPA diagnosis, we recorded the first AVTE and the time of occurrence. AVTE were classified as arterial or venous. Namely, venous events included deep venous thrombosis (DVT), pulmonary embolism, superficial venous thrombosis (SVT), and thrombosis at atypical locations, including ovarian, renal, splanchnic, jugular, retinal vein occlusion, and cerebral venous sinus thrombosis[17]. Arterial events included acute myocardial infarction, stroke, transient ischemic attack, acute ischemia of the upper and lower limbs, and arterial retinal occlusion.

Events related to acute myocardial infarction, stroke or pulmonary embolism were considered as potentially life-threatening.

For each patient, we calculated the accumulated person-time (person-years) as to the time elapsed from diagnosis until the first AVTE (when present) or the end of the follow-up.

### AVTE in the reference cohort

AVTE rates in EGPA patients where compared with those in the general population, using data from the Bruneck Study as a reference. The Bruneck Study is a prospective population-based study located in Northern Italy[18, 19]. In ascertaining AVTE outcomes, the participants’ self-reported medical history was carefully reviewed together with the medical records provided by general practitioners, Bruneck Hospital databases and death certificates. As in the EGPA cohort, time-to-event data in the Bruneck cohort was censored at the date of the first event, death, or end of follow-up, whichever was the earliest.

### Statistical analysis

No statistical sample size calculation was performed *a priori*.

Continuous variables are presented as median (interquartile range-IQR), while categorical variables as n (%).

Standardized event ratios (SERs) of AVTE in the EGPA cohort as compared to the reference Bruneck cohort were calculated. Time-to-event data of the Bruneck and EGPA cohorts were split by categories of age (i.e. <55 years, 55 to <65 years, 65 to <75 years, and ≥75 years) and the observed number of events in the EGPA cohort was compared to the expected event number if age-specific event rates had been identical to the Bruneck cohort. The 95% confidence intervals (CI) of the SERs were calculated as described by Rothman and Greenland [20].

Within the EGPA cohort, the risk of AVTE was estimated by fitting Cox regression models to calculate hazard ratios (HR) and related 95% CI and to derive Kaplan-Meier curves of AVTE. Covariates for model adjustments were selected from three areas: i) demographic characteristics (age, sex); ii) cardiovascular risk factors (previous AVTE, traditional atherogenic risk factors); and iii) EGPA-related variables (ANCA, BVAS, and eosinophils). To assess the robustness of our findings, a sensitivity analysis was performed excluding patients with history of AVTE prior to EGPA diagnosis. The analyses were performed using the software STATA version 14. Statistical significance was considered for p values <0.05.

## RESULTS

A total of 573 EGPA patients were included in the study. The main features recorded at time of EGPA diagnosis are described in **Table 1**. 297 patients (51.8%) were women; the median age at EGPA diagnosis was 55.3 years (IQR 44.8–64.0). Hypertension was the most frequent cardiovascular risk factor, followed by hypercholesterolemia. Regarding cardioprotective therapy, antihypertensive agents were the most employed drugs, followed by antiplatelet agents and statins. The most common EGPA manifestations included lower respiratory tract involvement (96.3%), ear-nose-throat involvement (79.4%), and peripheral neuropathy (63.2%). The median BVAS score at diagnosis was 12 (7-18); 130 patients had a five-factor score of zero, whereas 443 had a five-factor score ≥1. The results of ANCA testing were available for 485 patients. Of them, 243 (50.1%) were classified as ANCA-positive: 224 were ANCA-positive by IF, with most being positive for P-ANCA. Using ELISA, 215 patients were ANCA-positive; anti-myeloperoxidase (MPO) and anti-proteinase 3 (PR3) antibodies were respectively detected in 200 and 15 of them. Considering other laboratory parameters, the median values of C-reactive protein, erythrocyte sedimentation rate, eosinophil count and eosinophil cationic protein were respectively 5.2 mg/dL (2.1–10.4), 42.5 mm/h (23.0-61.0), 2680 U/mmc (1094.3–7600.0), and 52.0 µg/L (17.0–135.0). The median IgG4 level was 250.0 mg/dL (97.0–558.0), and the median IgE 400.0 U/mL (169.0–1000.0).

**Table 1:**
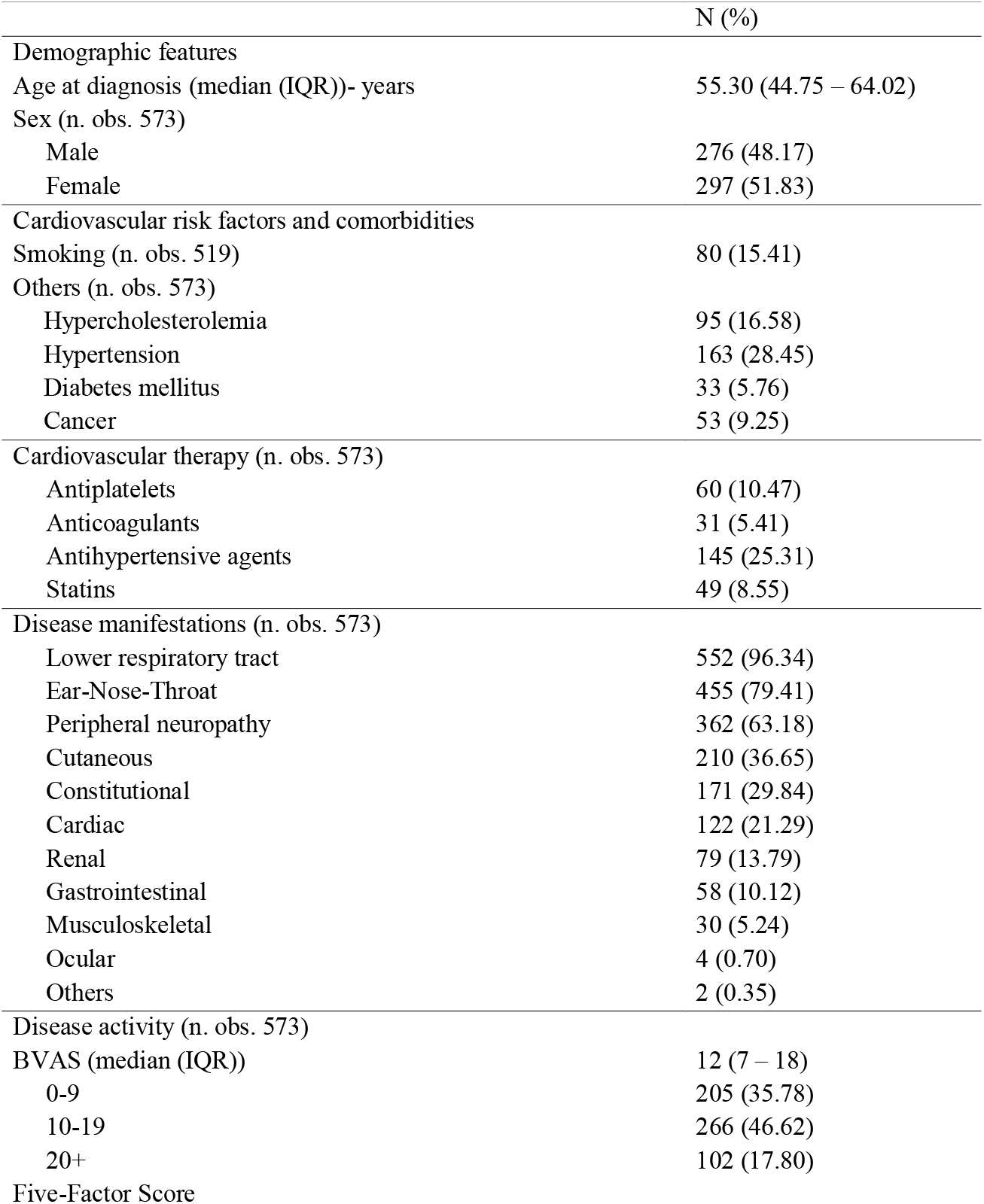
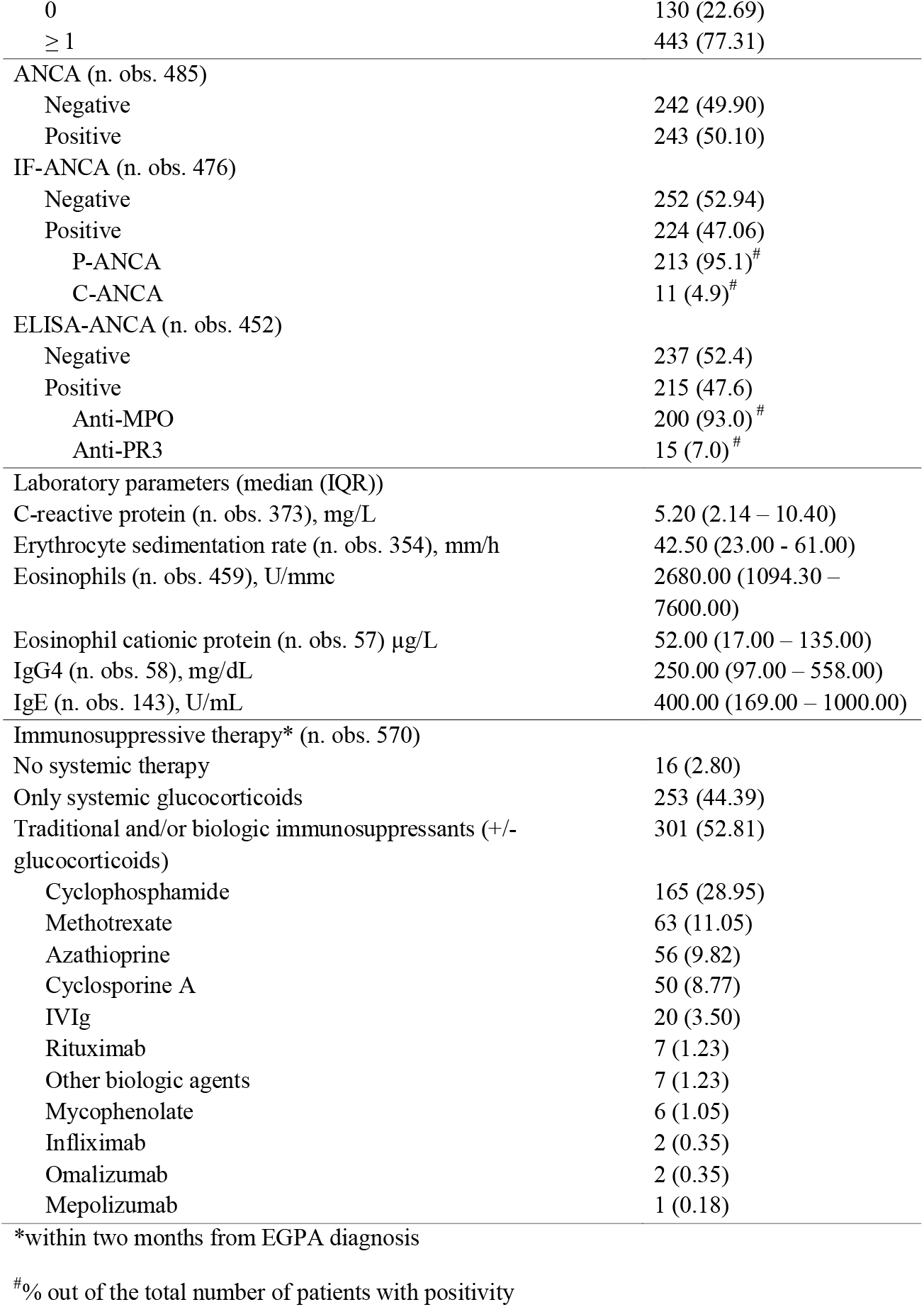
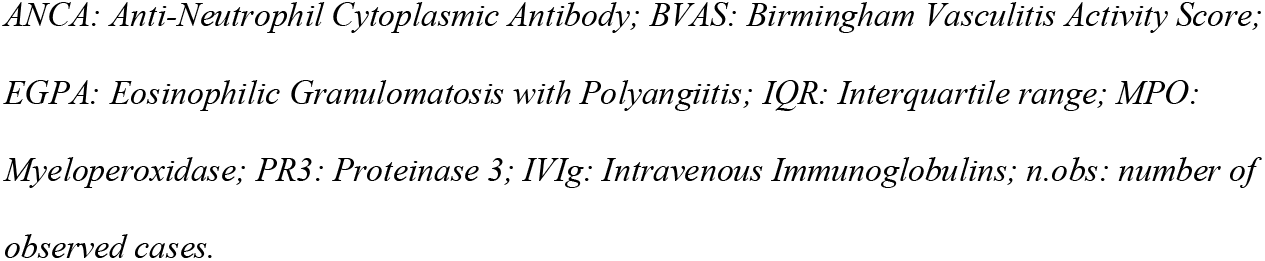
Demographic characteristics, cardiovascular risk factors, cardioprotective therapy and EGPA-related features at time of EGPA diagnosis.

|  | N (%) |
| --- | --- |
| Demographic features |  |
| Age at diagnosis (median (IQR))- years | 55.30 (44.75 – 64.02) |
| Sex (n. obs. 573) |  |
| Male | 276 (48.17) |
| Female | 297 (51.83) |
| Cardiovascular risk factors and comorbidities |  |
| Smoking (n. obs. 519) | 80 (15.41) |
| Others (n. obs. 573) |  |
| Hypercholesterolemia | 95 (16.58) |
| Hypertension | 163 (28.45) |
| Diabetes mellitus | 33 (5.76) |
| Cancer | 53 (9.25) |
| Cardiovascular therapy (n. obs. 573) |  |
| Antiplatelets | 60 (10.47) |
| Anticoagulants | 31 (5.41) |
| Antihypertensive agents | 145 (25.31) |
| Statins | 49 (8.55) |
| Disease manifestations (n. obs. 573) |  |
| Lower respiratory tract | 552 (96.34) |
| Ear-Nose-Throat | 455 (79.41) |
| Peripheral neuropathy | 362 (63.18) |
| Cutaneous | 210 (36.65) |
| Constitutional | 171 (29.84) |
| Cardiac | 122 (21.29) |
| Renal | 79 (13.79) |
| Gastrointestinal | 58 (10.12) |
| Musculoskeletal | 30 (5.24) |
| Ocular | 4 (0.70) |
| Others | 2 (0.35) |
| Disease activity (n. obs. 573) |  |
| BVAS (median (IQR)) | 12 (7 – 18) |
| 0-9 | 205 (35.78) |
| 10-19 | 266 (46.62) |
| 20+ | 102 (17.80) |
| Five-Factor Score |  |
| 0 | 130 (22.69) |
| ≥ 1 | 443 (77.31) |
| ANCA (n. obs. 485) |  |
| Negative | 242 (49.90) |
| Positive | 243 (50.10) |
| IF-ANCA (n. obs. 476) |  |
| Negative | 252 (52.94) |
| Positive | 224 (47.06) |
| P-ANCA | 213 (95.1)<sup>#</sup> |
| C-ANCA | 11 (4.9)<sup>#</sup> |
| ELISA-ANCA (n. obs. 452) |  |
| Negative | 237 (52.4) |
| Positive | 215 (47.6) |
| Anti-MPO | 200 (93.0)<sup>#</sup> |
| Anti-PR3 | 15 (7.0)<sup>#</sup> |
| Laboratory parameters (median (IQR)) |  |
| C-reactive protein (n. obs. 373), mg/L | 5.20 (2.14 – 10.40) |
| Erythrocyte sedimentation rate (n. obs. 354), mm/h | 42.50 (23.00 - 61.00) |
| Eosinophils (n. obs. 459), U/mmc | 2680.00 (1094.30 – 7600.00) |
| Eosinophil cationic protein (n. obs. 57) µg/L | 52.00 (17.00 – 135.00) |
| IgG4 (n. obs. 58), mg/dL | 250.00 (97.00 – 558.00) |
| IgE (n. obs. 143), U/mL | 400.00 (169.00 – 1000.00) |
| Immunosuppressive therapy* (n. obs. 570) |  |
| No systemic therapy | 16 (2.80) |
| Only systemic glucocorticoids | 253 (44.39) |
| Traditional and/or biologic immunosuppressants (+/- glucocorticoids) | 301 (52.81) |
| Cyclophosphamide | 165 (28.95) |
| Methotrexate | 63 (11.05) |
| Azathioprine | 56 (9.82) |
| Cyclosporine A | 50 (8.77) |
| IVIg | 20 (3.50) |
| Rituximab | 7 (1.23) |
| Other biologic agents | 7 (1.23) |
| Mycophenolate | 6 (1.05) |
| Infliximab | 2 (0.35) |
| Omalizumab | 2 (0.35) |
| Mepolizumab | 1 (0.18) |

\*within two months from EGPA diagnosis
<sup>#</sup>% out of the total number of patients with positivity
*ANCA: Anti-Neutrophil Cytoplasmic Antibody; BVAS: Birmingham Vasculitis Activity Score; EGPA: Eosinophilic Granulomatosis with Polyangiitis; IQR: Interquartile range; MPO: Myeloperoxidase; PR3: Proteinase 3; IVIg: Intravenous Immunoglobulins; n.obs: number of observed cases.*

After diagnosis, most patients (301, 52.5%) received traditional or biologic immunosuppressive agents, while only 44.4% were treated with systemic glucocorticoids alone. Only 2.8% of all patients did not receive immunosuppressive therapy within two months of diagnosis (including 1.6% who received delayed treatment, while 1.2% never received immunosuppression).

### AVTE occurrence and age-standardized rates

Overall, 129 EGPA patients (22.5%) had AVTE, including 70 (12%) prior to diagnosis and 75 (13%) following the diagnosis of EGPA [median follow-up 1677 days (IQR 663–3137)]. <u>AVTE prior to EGPA diagnosis.</u> Of the 70 patients who experienced AVTE before EGPA diagnosis (**Figure 1**), most did within the previous two years (n=41; 58%). Specifically, arterial events had occurred in 47 patients, whereas 18 patients had developed venous events and/or pulmonary embolism. In the remaining 5 patients, no information regarding the type of AVTE was available. Five patients had experienced two AVTEs each before diagnosis (both venous in two patients, both arterial in one, and one venous and one arterial in two. <u>AVTE after EGPA diagnosis.</u> After a median follow-up of 1677 days (IQR 663–3137) following EGPA diagnosis, 75 patients (13.1%) experienced at least one AVTE; the median time from the diagnosis of EGPA to the first AVTE was 577 days (IQR 81–2100). Ten events occurred at time of EGPA diagnosis, leading or contributing to identification of the underlying disease. Overall, forty-two out of the 75 events (56.0%) occurred in the two years following diagnosis (**Figure 1**), and >80% within the first six years. **Figure 2** shows the cumulative probability of a first AVTE after diagnosis in the whole cohort of patients. Thirty-two of these first AVTE were venous (19 DVT, 5 pulmonary embolism, 3 SVT, and 5 thrombosis at atypical locations, the latter including 2 Budd-Chiari syndromes, 2 retinal vein occlusions, 1 intracardiac thrombosis), whereas the remaining 43 were arterial (23 acute myocardial infarction, 10 stroke, 7 transient ischemic attack, 1 retinal artery occlusion, 1 acute upper limb ischemia, and 1 acute lower limb ischemia) (**Supplementary figure 1**). No information on the clinical prognosis and outcome following AVTE was available; however, 72/129 patients (55.9%) experienced events considered as potentially life-threatening (i.e., related to acute myocardial infarction, stroke, or pulmonary embolism).

**Figure 1.**
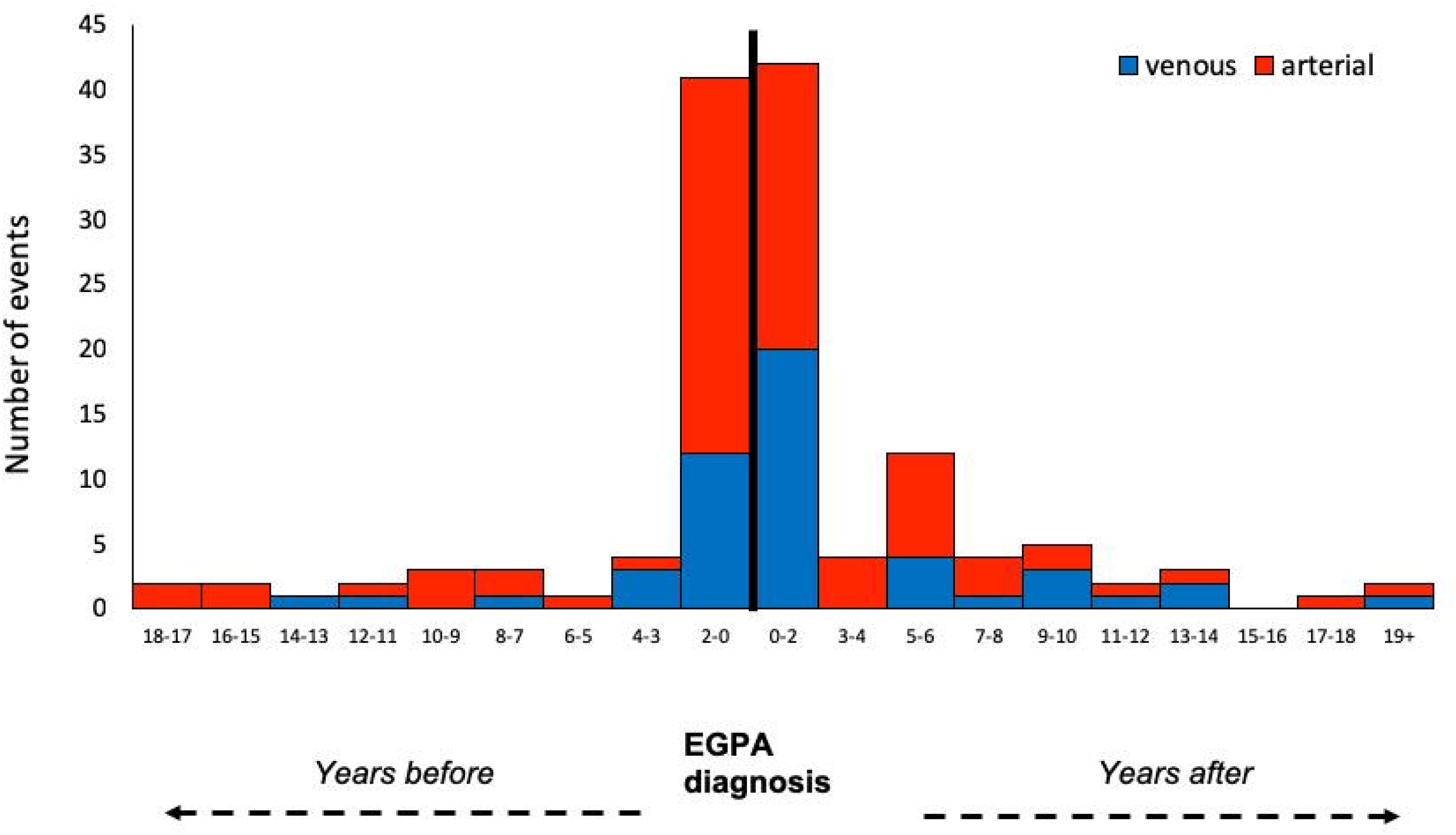
Time of occurrence of acute arterial and venous thromboembolic events before and after the diagnosis EGPA. *Events with missing data (i*.*e. type or timing of the event) are not reported. EGPA: Eosinophilic Granulomatosis with Polyangiitis*

**Figure 2.**
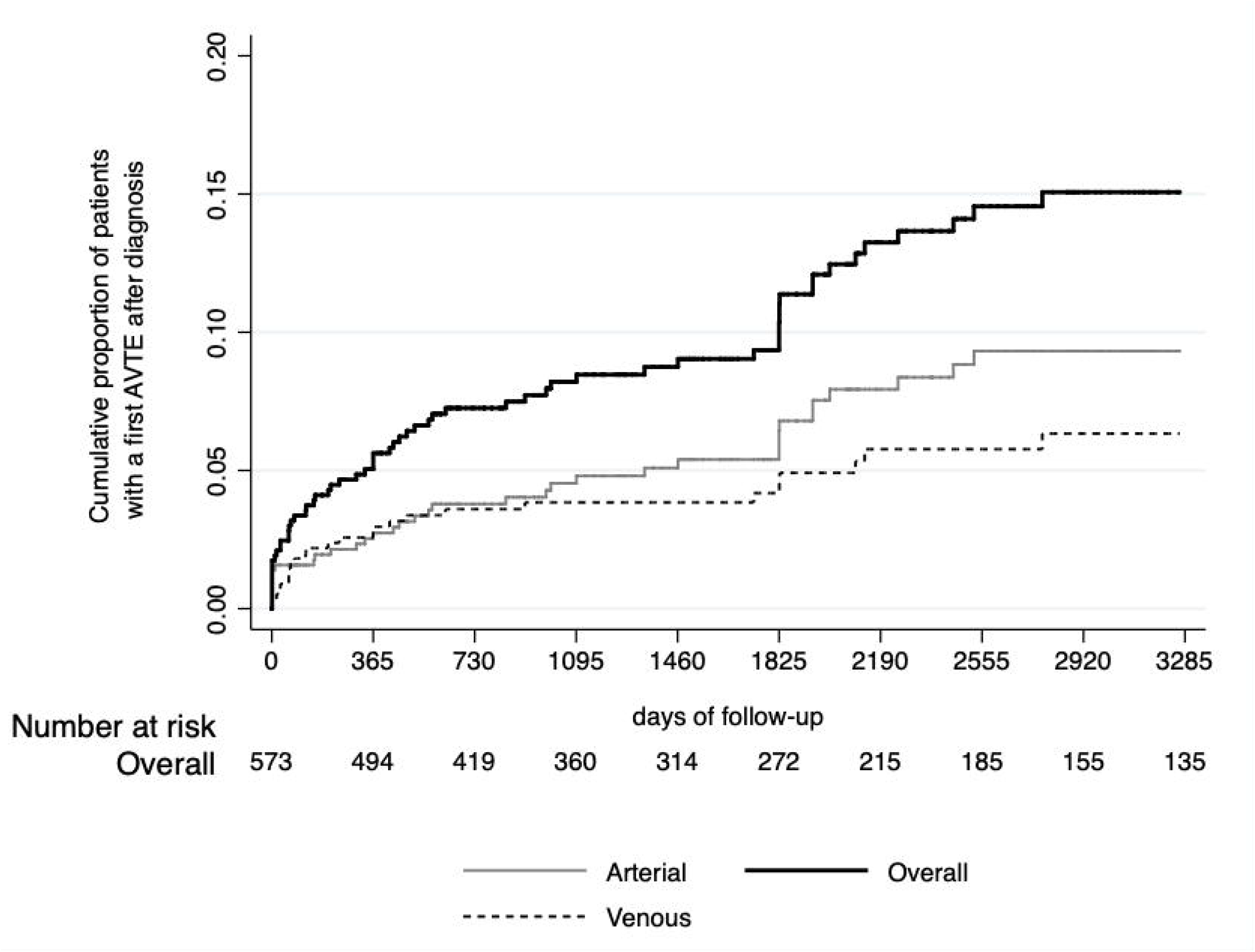
Kaplan-Meier curve of the occurrence of overall, arterial and venous thromboembolic events in the study cohort. *AVTE: arterial and venous thromboembolic events*

Age-standardized AVTE rates were calculated using a reference cohort of 933 individuals from the Bruneck Study (**Supplementary table 1**). In this reference cohort, 280 AVTE (226 arterial and 54 venous) were observed after a median follow-up of 20.9 years (10.3-25.6). Details on the observed AVTE are described in the **Supplementary table 2**.

The age-SER in EGPA patients as compared to the reference cohort was 2.10 (1.67-2.63; p<0.001) (**Table 2**). The highest SERs were observed for patients aged 55-65 years [3.24 (2.16-4.88)] or younger than 55 [3.12 (2.11-4.62)]. When stratifying the analysis according to the type of event, a SER of 1.64 (1.22-2.22) was found for arterial events (p<0.001); this rate was higher in EGPA patients younger than 65 years. With respect to venous events, the SER was 3.32 (2.35-4.70) (p<0.001); this increased risk in the EGPA cohort was statistically significant in all age classes.

**Table 2.** Rates of acute arterial and venous thromboembolic events standardized to the Bruneck cohort.

|  | Bruneck cohort,<br>rate per 1000<br>person-years | EGPA cohort,<br>observed no. of<br>events | EGPA<br>cohort,<br>expected<br>no. of<br>events | SER (95% CI) |
| --- | --- | --- | --- | --- |
| **All thromboembolic events** |  |  |  |  |
| Age category |  |  |  |  |
| <55 years | 5.7 (3.5-9.2) | 25 | 8.0 | 3.12 (2.11-4.62) |
| 55 to <65 years | 7.4 (5.3-10.4) | 23 | 7.1 | 3.24 (2.16-4.88) |
| 65 to <75 years | 16.5 (13.3-20.5) | 13 | 12.5 | 1.04 (0.60-1.78) |
| ≥75 years | 34.4 (29.3-40.4) | 14 | 8.2 | 1.72 (1.02-2.90) |
| Overall |  | 75 | 35.8 | 2.10 (1.67-2.63)<br>P<0.001 |
| **Arterial events** |  |  |  |  |
| Age category |  |  |  |  |
| <55 years | 2.5 (1.2-5.2) | 17 | 3.5 | 4.86 (3.02-7.81) |
| 55 to <65 years | 5.8 (4.0-8.6) | 13 | 5.6 | 2.33 (1.35-4.01) |
| 65 to <75 years | 13.3 (10.5-16.9) | 5 | 10.1 | 0.49 (0.21-1.19) |
| ≥75 years | 29.3 (24.6-34.9) | 8 | 6.9 | 1.15 (0.58-2.31) |
| Overall |  | 43 | 26.1 | 1.64 (1.22-2.22)<br>P<0.001 |
| **Venous events** |  |  |  |  |
| Age category |  |  |  |  |
| <55 years | 3.2 (1.7-6.1) | 8 | 4.5 | 1.78 (0.89-3.55) |
| 55 to <65 years | 1.6 (0.7-3.3) | 10 | 1.5 | 6.65 (3.58-12.36) |
| 65 to <75 years | 3.2 (1.9-5.2) | 8 | 2.4 | 3.31 (1.65-6.62) |
| ≥75 years | 5.1 (3.4-7.8) | 6 | 1.2 | 4.95 (2.23-11.03) |
| Overall |  | 32 | 9.6 | 3.32 (2.35-4.70)<br>P<0.001 |

*CI: confidence interval; SER standardized event ratio.*

### AVTE predictors after EGPA diagnosis

**Figure 3a** shows adjusted HRs of AVTE based on different demographic and clinical features recorded at diagnosis. A significantly higher risk of AVTE was found in patients with a history of previous AVTE (HR 2.06 [95%CI 1.15-3.67], p=0.015) and in patients with BVAS ≥ 20 at diagnosis [HR 2.02 (95%CI 1.03–3.96) as compared with patients with BVAS 0-9, p=0.041], while no association was found between the five-factor score and the development of AVTE. Furthermore, no association was found with ANCA positivity, sex, age at diagnosis and presence of classic cardiovascular risk factors. Of note, the association between BVAS and AVTE was confirmed also when limiting the analyses to patients with no history of AVTE before EGPA diagnosis (**Supplementary table 3**). On the other hand, baseline levels of eosinophils as well as of C-reactive protein, erythrocyte sedimentation rate, IgG4 and IgE levels did not influence the risk of AVTE (data not shown). The influence of eosinophil cationic protein levels on the risk of AVTE was not assessed, due to the high proportion of missing data.

**Figure 3.**
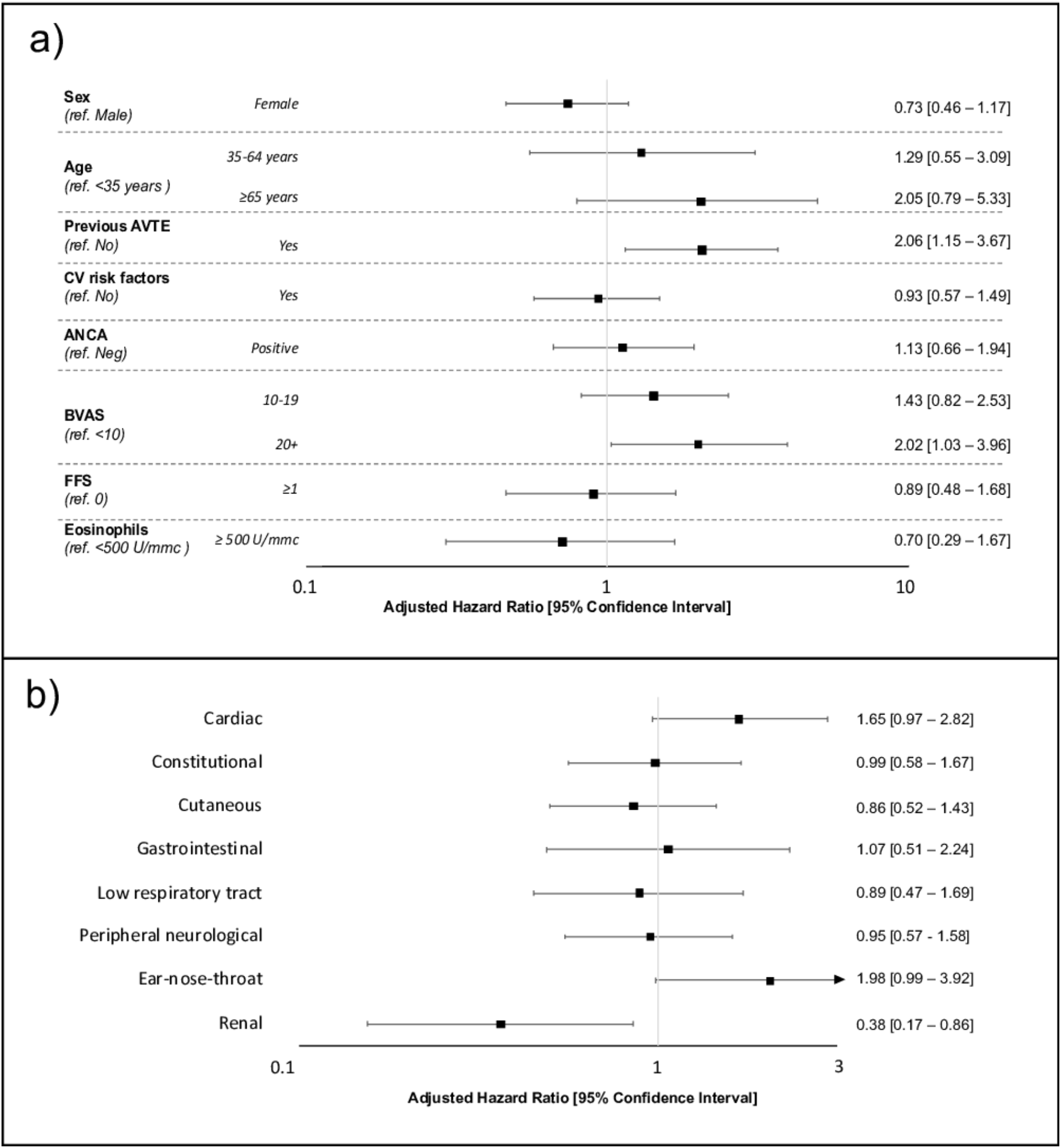
Adjusted risks of acute arterial and venous thromboembolic events (AVTE), according **a)** to demographic, clinical and EGPA-related features, and **b)** to specific EGPA manifestations. *Risks are adjusted for age, sex, previous AVTE, ANCA, BVAS, eosinophils, and presence of cardiovascular risk factors*. *ANCA: Anti-Neutrophil Cytoplasmic Antibody; AVTE: arterial and venous thromboembolic events; BVAS: Birmingham Vasculitis Activity Score; CV: Cardiovascular; FFS: Five-Factor Score*.

Regarding disease manifestations (**Figure 3b**), a slightly higher, but of borderline statistical significance, risk of AVTE was found for patients with ear-nose-throat or cardiac involvement [HR 1.98 (95%CI 0.99–3.92]) and 1.65 (95%CI 0.97-2.82), respectively], whereas renal involvement was associated with a lower risk of AVTE [HR 0.38 (95%CI 0.17–0.86), p=0.020]. None of the other clinical manifestations significantly influenced the risk of AVTE.

### Pharmacological treatments and AVTE occurrence

We finally investigated the role of immunosuppressive and cardiovascular therapy at EGPA diagnosis on the risk of AVTE occurrence. The 16 patients who did not receive any immunomodulating therapy within the first two months of diagnosis had a significantly higher risk of AVTE as compared with patients receiving systemic glucocorticoids [HR 3.67 (1.37– 9.89), p=0.010], whereas risk was comparable in patients treated with immunosuppressants as compared with systemic glucocorticoids alone (data not shown). Of note, the association between the lack of immunosuppression within 2 months of diagnosis and risk of AVTE was confirmed also when considering only patients with no history of AVTE prior to EGPA diagnosis (data not shown).

With respect to the ongoing cardioprotective treatments at the time of diagnosis, no association was found between antiplatelet, anticoagulant or statin therapy and the risk of AVTE (adjusted HR of 0.99 [95%CI 0.98–1.01], p=0.985 for antiplatelets, 0.35 [95%CI 0.09– 1.57], p=0.176 for anticoagulants, and 0.96 [95%CI 0.40–2.26], p=0.918 for statins).

## DISCUSSION

In the present study, presenting the largest EGPA cohort reported to date, we observed a significant increase in age standardized risk of arterial and venous acute thromboembolic events associated with the disease compared to the general population. Overall, 23% of 573 patients developed AVTE, both prior and following the diagnosis of EGPA, half of them being potentially life threatening and representing the reason for initial diagnosis of the underlying disease in 10 cases.

Notably, the majority of these events clustered in the two years before and the two years after the diagnosis and peaked in the periods of highest disease activity. Conversely, classic cardiovascular risk factors had no impact on risk and, specifically, EGPA was associated with increased rates of arterial events compared to the reference population in spite of a more favorable cardiovascular risk profile.

The fact that active inflammation, rather than traditional mechanisms of thrombosis, promotes AVTE in EGPA, is strongly supported by the observed effects of treatment. The small subset of patients who did not receive systemic glucocorticoids or immunosuppressants during the first two months after EGPA diagnosis were at distinctively greater risk of AVTEs compared to treated patients. Conversely, lack of both anticoagulant and antiplatelet agent use did not seem to confer any added risk. These findings lend further support to the view that AVTEs are directly determined by active EGPA vascular involvement, and may be prevented by adequate and timely immunosuppression[21–23]. Although these findings may also reflect the fact that anticoagulant/antiplatelet agents are more likely prescribed to the most severe patients, thus representing a bias in our analysis, these novel observations warrant further investigation in the field.

The results of the present study are consistent with the notion that EGPA evolves through different phases, namely an allergic phase which usually lasts for years, a eosinophilic phase of shorter duration, and a rapidly evolving vasculitic phase[1]. This sequence of disease stages suggests that eosinophilic inflammation is already active before the full-blown disease manifestations develop and may play a decisive role in the development of AVTEs soon before and after the initial diagnosis, plausibly as a reflection of uncontrolled active disease. Indeed, we found that patients younger than 65 years had the highest SER of AVTE. Considering that in our cohort around 75% of patients were diagnoses at <65 years of age, this result further corroborates our hypothesis that the high disease activity around diagnosis probably influences the AVTE risk.

In our cohort, the majority of AVTEs were arterial and mainly included acute myocardial infarction and stroke. Most published studies focused on the association between AAVs and venous thromboembolism[5, 6, 23], while limited evidence supports an association with arterial manifestations[3, 4, 24]. Venous thromboembolic events, although less frequent than arterial events, were associated with the highest SER as compared to the reference population. As expected, most were lower limb DVT or pulmonary embolism, although a few patients developed thrombosis at atypical sites. In a retrospective analysis of the French Vasculitis Study Group including 232 EGPA patients, 8.2% developed venous thromboembolic events after a mean follow-up of 58 months. Interestingly, most events occurred between 3 months before and 6 months after diagnosis[6]. A Russian study including 69 EGPA cases reported that 5 of them (7.2%) developed venous thromboembolism, all during the first year after diagnosis[25]. With regard to the other AAVs, a particular increase in risk of DVT[7, 26] and pulmonary embolism [7] was observed during active disease[27].

Finally, we identified several predictors on AVTE in EGPA. Patients who had experienced AVTEs before diagnosis were at higher risk of developing additional events following diagnosis, likely related to pre-existing vascular injury and individual susceptibility[28]. This, close clinical monitoring is required in secondary prevention for these patients[6, 26]. Furthermore, while ear-nose-throat and cardiac manifestations seemed associated with an increased risk of AVTE, renal involvement did not. Whether these discrepancies reflect the diverse biology of the two main clinical subsets of EGPA, *i*.*e*. vasculitic vs eosinophilic (with the latter being more prone to AVTE), is unclear. The lack of association between ANCA status and AVTE does not seem to corroborate this hypothesis.

Our study has other limitations, some of which are inherent to retrospective studies. First, as AVTEs diagnoses were retrospectively captured from medical charts, misclassification of the events might have occurred. Second, the role of some heterogeneity in clinical management due to the long-term study period cannot be excluded. Finally, no information on the clinical prognosis and outcome following AVTE was available. Despite these limitations, our study finds its strengths in its large sample size, the long follow-up, and the presence of a reference population.

In conclusion, our results show a higher risk of both acute AVTE events in patients with EGPA as compared to a reference population, particularly around the time of EGPA diagnosis and in those without immunosuppressive treatments. Notably, the occurrence of venous events was stably high throughout the whole duration of the disease, suggesting the need of a close and long-term monitoring particularly for such events. Based on our data EGPA seems to represent a cardiovascular risk factor independently from the patient’s demographic and clinical features, and AVTE risk is particularly increased in patients with previous AVTE and high disease activity. These findings also suggest that the occurrence of unexplained AVTE in healthy subjects should be investigated in order to exclude an underlying eosinophilic condition, and particularly EGPA.

## Data Availability

Deidentified participant data are available upon reasonable request, to be sent by email to the corresponding author.

## ACKNOWLEDGEMENTS

All people who contributed to this manuscript are listed as authors or collaborators.

## **Collaborators of the EGPA Italian Consortium

Elena Bargagli (Department of Medicine, Surgery and Neurosciences, Respiratory Diseases and Lung Transplantation, Regional Referral Centre for Sarcoidosis and ILD, Siena University, Siena), Matteo Becatti (Department of Experimental and Clinical Biomedical Sciences “Mario Serio”, University of Firenze, Firenze, Italy), Mirko Beccalli (Internal Medicine, Clinical Immunology and Translational Medicine Unit, IRCCS Ospedale Policlinico San Martino, Genoa, Italy), Federica Bello (Department of Experimental and Clinical Medicine, University of Firenze, Italy), Francesco Bozzao (Rheumatology Unit, Medicina Clinica, Cattinara Teaching Hospital (ASUITS) Trieste)), Valentina Canti (Unit of Immunology, Rheumatology, Allergy and Rare Diseases (UnIRAR), IRCCS-San Raffaele Scientific Institute, Milan, Italy), Matthias A. Cassia (ASST Santi Paolo e Carlo and the University of Milan, Milan, Italy), Giulia Cassone (Clinical and Experimental Medicine PhD Program, Azienda USL-IRCCS di Reggio Emilia and Università di Modena and Reggio Emilia, Italy), Mariagrazia Catanoso (Rheumatology Unit, Department of Specialistic Medicine, Azienda USL-IRCCS di Reggio Emilia, Italy), Fulvia Chieco-Bianchi (Respiratory Pathophysiology Division, University Hospital of Padova, Italy), Roberta Clari (Nephrology, Dialysis and Renal Transplant Division, Department of Medical Sciences, “Città della Salute e della Scienza di Torino” University Hospital, University of Turin, Italy), Laura Coladonato (Rheumatology Unit, Department of Emergency and Organ Transplantation (DETO), Polyclinic Hospital, University of Bari, Italy), Maria De Santis (Division of Rheumatology and Clinical Immunology, Humanitas Clinical and Research Center - IRCCS, Rozzano, Milan, Italy), Gerardo Di Scala (Department of Experimental and Clinical Medicine, University of Firenze, Italy), Filippo Fagni (Department of Experimental and Clinical Medicine, University of Firenze, Italy), Paride Fenaroli (Nephrology Unit, Parma University Hospital, Parma, Italy), Claudia Fiorillo (Department of Experimental and Clinical Biomedical Sciences “Mario Serio”, University of Firenze, Firenze, Italy), Alberto Floris (Rheumatology, Department of Medical Sciences and Public Health, Univerisity Clinic, Cagliari, Italy), Marco Fornaro (Rheumatology Unit, Department of Emergency and Organ Transplantation (DETO), Polyclinic Hospital, University of Bari, Italy), Elena Galli (University of Modena and Reggio Emilia, Modena, Italy), MD, Elena Generali (Division of Rheumatology and Clinical Immunology, Humanitas Clinical and Research Center, IRCCS, Rozzano, Milan, Italy), Marica Giliberti (Department of Emergency and Organ Transplantation, Nephrology, Dialysis and Transplant Unit, University of Bari “Aldo Moro”, Bari, Italy), Nancy Lascaro (Rheumatology Institute of Lucania (IRel) and the Rheumatology Department of Lucania, San Carlo Hospital of Potenza and Madonna delle Grazie Hospital of Matera, Potenza and Matera, Italy), Ilaria Leccese (Rheumatology Unit, Department of Clinical Internal, Anesthesiological and Cardiovascular Sciences, Sapienza University of Rome, Rome, Italy), Irene Mattioli (Department of Experimental and Clinical Medicine, University of Firenze, Italy), Bianca Olivieri (Department of Medicine, Università degli Studi di Verona, Italy), Nicola Osti (Department of Medicine, Università degli Studi di Verona, Italy), Francesco Peyronel (Nephrology Unit, University Hospital, Parma, Italy), Massimo Radin (Department of Clinical and Biological Sciences, University of Turin, Italy), Giulia Righetti (Rheumatology Unit, Department of Emergency and Organ Transplantation (DETO), Polyclinic Hospital, University of Bari, Italy), Stefano Salvati (Vita-Salute San Raffaele University, Milan, Italy), Elena Silvestri (Department of Experimental and Clinical Medicine, University of Firenze, Italy), Nicola Susca (Department of Biomedical Sciences and Human Oncology, Unit of Internal Medicine “Guido Baccelli”, University of Bari “Aldo Moro” Medical School, Bari, Italy), Carlo Tamburini (Department of Experimental and Clinical Medicine, University of Firenze, Italy; SOD Interdisciplinary Internal Medicine, Center for Autoimmune Systemic Diseases-Behçet Center and Lupus Clinic-AOU Careggi Hospital of Florence, Florence, Italy), Giusy Taurisano (Department of Experimental and Clinical Medicine, University of Firenze, Italy), Barbara Trezzi (Department of Medicine and Surgery, University of Milano – Bicocca and Nephrology Unit, ASST-Monza, Milan/Monza, Italy), Giorgio Trivioli (Nephrology and Dialysis Unit, Meyer Children’s Hospital; Department of Experimental and Clinical Biomedical Sciences “Mario Serio”, University of Firenze, Firenze, Italy).

Giacomo Emmi, Augusto Vaglio, Domenico Prisco, Iacopo Olivotto, Renato Alberto Sinico, Franco Schiavon, Sara Monti, Enrica Paola Bozzolo, Franco Franceschini, Marcello Govoni, Claudio Lunardi, Giuseppe Guida, Giuseppe Lopalco, Giuseppe Paolazzi, Angelo Vacca, Gina Gregorini, Pietro Leccese, Matteo Piga, Fabrizio Conti, Paolo Fraticelli, Luca Quartuccio, Federico Alberici, Carlo Salvarani, Silvano Bettio, Simone Negrini, Carlo Selmi, Savino Sciascia, Gabriella Moroni Loredana Colla, and Carlo Manno conceived the study.

Giacomo Emmi, Augusto Vaglio, Domenico Prisco, Peter Willeit and Alessandra Bettiol designed the study.

Maria Letizia Urban, Alfredo Vannacci, Maria Rosa Pozzi, Paolo Fabbrini, Stefano Polti, Mara Felicetti, Maria Rita Marchi, Roberto Padoan, Paolo Delvino, Roberto Caporali, Carlomaurizio Montecucco, Lorenzo Dagna, Adriana Cariddi, Paola Toniati, Silvia Tamanini, Federica Furini, Alessandra Bortoluzzi, Elisa Tinazzi, Lorenzo Delfino, Iuliana Badiu, Giovanni Rolla, Vincenzo Venerito, Florenzo Iannone, Alvise Berti, Roberto Bortolotti, Vito Racanelli, Guido Jeannin, Angela Padula, Alberto Cauli, Roberta Priori, Armando Gabrielli, Milena Bond, Martina Tedesco, Giulia Pazzola, Paola Tomietto, Marco Pellecchio, Chiara Marvisi, Federica Maritati, Alessandra Palmisano, Christian Dejaco, Johann Willeit, Stefan and Kiechl acquired the data, together will all authors’ collaborators. Alessandra Bettiol and Peter Willeit performed the statistical analysis. Giacomo Emmi, Augusto Vaglio, and Domenico Prisco interpreted the results. Alessandra Bettiol wrote the manuscript, assisted by Giacomo Emmi and Augusto Vaglio. All authors and co-authors critically revised the manuscript, approved the final version of this manuscript, and agreed to be accountable for all aspects of the work, ensuring that questions related to the accuracy or integrity of any part of the work are appropriately investigated and resolved.

