## Supplementary figure1; Supplementary tables 1-3 for "Risk of acute arterial and venous thromboembolic events in Eosinophilic Granulomatosis with Polyangiitis (Churg-Strauss syndrome)": SUPPLEMENTAL MATERIAL_Pre-print.docx

**Supplementary figure 1.** Types of acute arterial and venous events occurred in the cohort of EGPA patients after diagnosis. In bold are events with frequency of 5 or more.

EGPA: Eosinophilic Granulomatosis with Polyangiitis

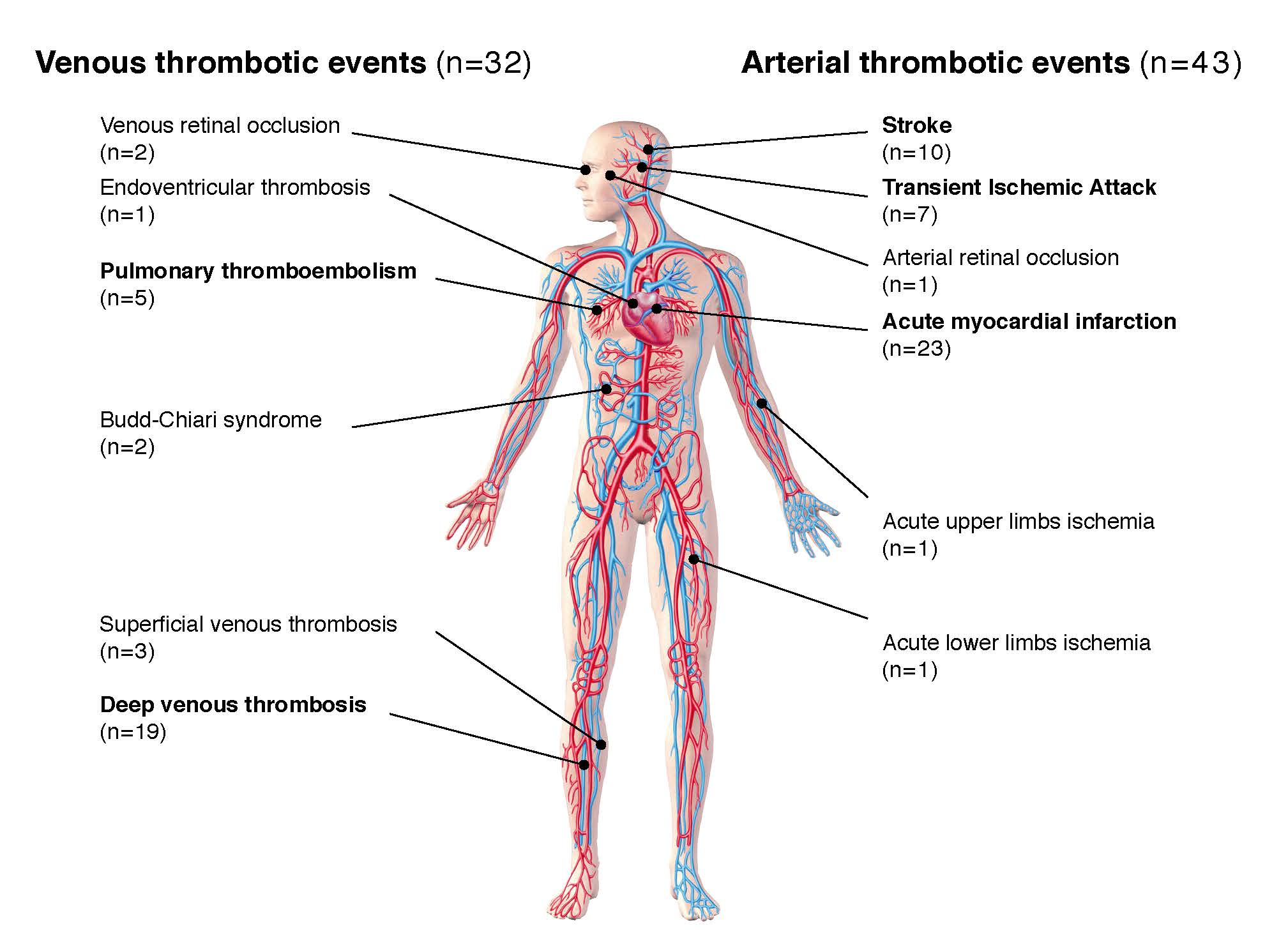

**Supplementary table 1.** Descriptive summary of the Bruneck cohort (n=933).

|  | N (% out of 933) |
| --- | --- |
| Year of recruitment of the cohort | 1990 |
| **Demographic features** |  |
| Female sex | 459 (49⋅2%) |
| Median age at entry (IQR), years | 59⋅2 (49⋅1-68⋅8) |
| **Cardiovascular risk factors and comorbidities** |  |
| Smoking | 228 (24⋅44) |
| Others |  |
| Hypercholesterolemia | 658 (70⋅53) |
| Hypertension | 597 (63⋅99) |
| Diabetes mellitus | 77 (8⋅25) |
| Cancer | 33 (3⋅54) |
| **Cardiovascular therapy** |  |
| Antiplatelets | 31 (3⋅32) |
| Anticoagulants | 3 (0⋅32) |
| Antihypertensive agents | 172 (18⋅44) |
| Statins | 16 (1⋅71) |
| **Occurrence of AVTE** |  |
| Median age at censoring (IQR), years | 77⋅2 (70⋅3-83⋅9) |
| Median duration of follow-up (IQR), years | 20⋅9 (10⋅3-25⋅6) |
| Thromboembolic event rate (95% CI), per 1000 person-years | 16⋅8 (15⋅0-18⋅9) |
| Venous thromboembolic event rate (95% CI), per 1000 person-years | 3⋅2 (2⋅5-4⋅2) |
| Arterial thromboembolic event rate (95% CI), per 1000 person-years | 13⋅6 (11⋅9-15⋅5) |

AVTE: arterial and venous thromboembolic events; CI: confidence interval; IQR: interquartile range*.*

**Supplementary table 2.** Number of acute venous and arterial thromboembolic events in the Bruneck cohort.

|  | Bruneck cohort (n=933) |
| --- | --- |
| **Venous thromboembolic events** |  |
| No. of events | 54 |
| Deep vein thrombosis | 33 |
| Pulmonary embolism | 13 |
| Superficial vein thrombosis | 1 |
| Budd-Chiari syndrome | 0 |
| Retinal vein occlusion | 7 |
| Intracardiac thrombosis | 0 |
| **Arterial thromboembolic events** |  |
| No. of events | 226 |
| Acute myocardial infarction | 73 |
| Stroke | 105 |
| Transient ischaemic attacks | 45 |
| Retinal artery occlusion | 0 |
| Acute upper limb ischaemia | 0 |
| Acute lower limb ischaemia | 3 |
| **All thromboembolic events** |  |
| No. of events | 280 |

**Supplementary table 3.** Adjusted Hazard Ratio [95% Confidence Intervals] of arterial and venous thromboembolic events (AVTE) among patients with no history of previous AVTE.

|  | **Adjusted HR [95% CI] #** | **p-value** |
| --- | --- | --- |
| Sex |  |  |
| Male | Ref. |  |
| Female | 0⋅75 [0⋅44 – 1⋅27] | 0⋅281 |
| Age |  |  |
| <35 years | Ref. |  |
| 35-65 years | 1⋅44 [0⋅56 – 3⋅71] | 0⋅451 |
| ≥65 years | 2⋅67 [0⋅93 – 7⋅67] | 0⋅069 |
| Presence of CV risk factors |  |  |
| No | Ref. |  |
| Yes | 0⋅78 [0⋅45 – 1⋅32] | 0⋅352 |
| ANCA |  |  |
| Negative | Ref. |  |
| Positive | 0⋅94 [0⋅51 – 1⋅74] | 0⋅846 |
| *Missing* | 1⋅29 [0⋅62 – 2⋅70] | 0⋅497 |
| BVAS |  |  |
| 0-9 | Ref. |  |
| 10-19 | 1⋅29 [0⋅67 – 2⋅44] | 0⋅2440 |
| 20+ | 2⋅22 [1⋅04 – 4⋅70] | 0⋅038* |
| Five-Factor Score |  |  |
| 0 | Ref. |  |
| ≥1 | 0⋅76 [0⋅38 – 1⋅52] | 0⋅438 |
| Eosinophils |  |  |
| <500 cells/UL | Ref. |  |
| >=500 cells/UL | 0⋅52 [0⋅21 – 1⋅29] | 0⋅159 |
| *Missing* | 0⋅71 [0⋅26 – 1⋅92] | 0⋅505 |
| CRP |  |  |
| <0.9 mg/dL | Ref. |  |
| ≥0.9 mg/dL | 1⋅06 [0⋅33 – 3⋅33] | 0⋅927 |
| *Missing* | 2⋅00 [0⋅61 – 6⋅57] | 0⋅251 |
| ESR |  |  |
| ≤25 mm/h | Ref. |  |
| >25 mm/h | 0⋅56 [0⋅25 – 1⋅28] | 0⋅171 |
| *Missing* | 1⋅15 [0⋅51 – 2⋅59] | 0⋅729 |
| Immunosuppressive therapy |  |  |
| Only systemic steroids | Ref. |  |
| No systemic therapy | 3⋅08 [0⋅98 – 9⋅71] | 0⋅054 |
| Traditional and/or biologic  immunosuppressants (+/- steroids) | 0⋅89 [0⋅51 – 1⋅55] | 0⋅671 |

\# adjusted for age, sex, ANCA, BVAS, eosinophils, and presence of CV risk factors.

*ANCA: Anti-Neutrophil Cytoplasmic Antibody; BVAS: Birmingham Vasculitis Activity Score; CV: Cardiovascular; CRP: C-Reactive Protein; ESR: Erythrocyte Sedimentation Rate*

**statistically significant for p-value<0*⋅*05*
